# Optimized infection control practices augment the robust protective effect of vaccination for ESRD patients during a hemodialysis facility SARS-CoV-2 outbreak

**DOI:** 10.1101/2022.03.18.22272356

**Authors:** Megan E. Meller, Bridget L. Pfaff, Andrew J. Borgert, Craig S. Richmond, Deena M. Athas, Paraic A. Kenny, Arick P. Sabin

## Abstract

**Background:** While dialysis patients are at greater risk of serious SARS-CoV-2 complications, stringent infection prevention measures can help mitigate the risk of infection and transmission within dialysis facilities. We describe an outbreak of 14 cases diagnosed in a 13-day period in the second quarter of 2021 in a hospital-based ESRD facility, and our coordinated use of epidemiology, viral genome sequencing, and infection control practices to quickly end the cycle of transmission.

**Methods:** Symptomatic patients and staff members were diagnosed via RT-PCR tests. Facility-wide screening was conducted using rapid SARS-CoV-2 antigen tests. SARS-CoV-2 genome sequences were obtained from residual diagnostic PCR specimens.

**Results:** Of the 106 patients who received dialysis in the facility, 10 were diagnosed with SARS-CoV-2 infection, as was one patient support person. Of three positive staff members, two were unvaccinated and had provided care for six and four of the affected patients, respectively. Sequencing demonstrated that all the cases in the cluster shared an identical B.1.1.7./Alpha substrain. Attack rates were greatest among unvaccinated patients and staff. Vaccine effectiveness was 88% among patients.

**Conclusions:** Prompt recognition of an infection cluster and rapid intervention efforts successfully ended the outbreak. Alongside consistent adherence to core infection prevention measures, vaccination was highly effective in reducing disease incidence and morbidity in this vulnerable population.

## INTRODUCTION

SARS-CoV-2 infection poses a particularly acute risk to end stage renal disease (ESRD) patients. During the first seven months of the pandemic, it is estimated that excess deaths in this population were 8.7-12.9 per 1000 patients [1]. In a study of ESRD patients with COVID-19, 67% required emergency department or inpatient care, and the mortality rate exceeded 20% [2]. Coupled with their higher risk of adverse outcomes and death, dialysis patients have weaker responses to the vaccine overall, with 22% of fully vaccinated individuals having either absent or an attenuated antibody response [3], and antibody levels among those who do mount responses are markedly lower than in non-dialysis controls [4].

Dialysis facilities have adopted several interventions to combat SARS-CoV-2 and still maintain the ability to perform life-sustaining therapy despite widespread community transmission. Facility control plans have been implemented to include protection for this high-risk population in a congregate setting. Some of these core interventions include masking, physical distancing, rapid case identification and vigorous vaccination programs. Facilities are also encouraged to partner jointly with local public health and the Centers for Disease Control (CDC) in response to potential outbreaks.

Rapid turnaround viral genome sequencing aids differentiation of outbreaks from pseudo-outbreaks by way of comparing viral genomes of contemporaneous cases to those circulating elsewhere in the community. Distinguishing these scenarios can usefully inform the appropriate facility response strategy and conserve resources that would otherwise be expended needlessly on pseudo-outbreaks. Using this approach, our group previously analyzed a cluster of 5 hemodialysis facility cases with all the epidemiological hallmarks of nosocomial transmission, but which real-time genetic analysis confirmed were wholly unrelated, providing an example of the infection control conundrum that can arise during a community surge [5].

Here we describe a series of 14 cases occurring within a 13-day period in a hospital-based ESRD facility in Southwest Wisconsin. With far higher patient (88%) and staff (77%) vaccination rates (Table 1) than reported ESRD facility averages in Wisconsin at that time (70% and 50%, respectively [6]), this analysis also underscores the persistent outbreak risk remaining in a setting with strong, albeit incomplete, vaccine coverage in commingled persons. Case epidemiology, facility-wide surveillance, and genetic analysis to elucidate near real-time transmission dynamics were integrated to influence enhanced infection control recommendations and decisively end the outbreak.

**Table 1.** Vaccination Status of ESRD facility patients and staff.

| Table 2: Vaccination Status Summary |  |  |
| --- | --- | --- |
|  | Patients (N=106) | Staff (N=47) |
| Unvaccinated, N (%) | 10 (9%) | 11 (23%) |
| Partially Vaccinated, N (%) | 3 (3%) | 0 |
| Fully Vaccinated, N (%) | 93 (88%) | 36 (77%) |

## METHODS

### Epidemiological Investigation

An epidemiological investigation was initiated in response to a cluster of SARS-CoV-2 infections in a 32-bed outpatient dialysis facility. Information on patient symptoms, clinical outcomes, individual interactions at the facility, dialysis schedule, transportation, and vaccine administration were collected. Infection prevention and control (IPC) assessments were conducted to identify IPC breaches that may have contributed to the outbreak. Local and state public health officials were consulted.

### Case Definition and Identification

All symptomatic patients and staff members were diagnosed via RT-PCR using nasopharyngeal specimens. A confirmed case of SARS-CoV-2 was defined as having a newly positive RT-PCR during the outbreak span. A probable case of SARS-CoV-2 was defined as having a positive SARS-CoV-2 antigen test. Cases were given an identifier starting with either “P” (patients) or “S” (staff).

Facility-wide surveillance testing was performed by the Infection Control department based on CMS’s guidance for surveillance testing in long term care [7]. Patients and staff members were tested for SARS-CoV-2 onsite at the facility with the Abbot Binax-NOW antigen kit, with testing occurring twice a week to account for the alternating dialysis shifts (schedule A and schedule B). Patients were screened at their treatment station. Staff members were screened in a conference room located on site. Staff participation in surveillance testing was mandatory. Two staff members refused to participate and were not allowed to return to work until the outbreak was declared over.

Patient participation in surveillance testing was optional; patients were informed that if they refused to participate or to wear a mask, they would be treated as SARS-CoV-2 positive and placed into isolation for dialysis treatment. This decision was supported by local public health and reflected practices used contemporaneously in long-term care settings. Only one patient in the facility refused to participate during the span of the outbreak.

### SARS-CoV-2 Sequencing and Analysis

cDNA was generated from residual RNA from diagnostic specimens using ProtoScript II (New England Biolabs, Ipswich, MA). The Ion AmpliSeq SARS-CoV-2 Panel (Thermo-Fisher, Waltham, MA) was used to amplify 237 viral specific targets encompassing the complete viral genome. Libraries were sequenced and analyzed as we have previously described [5, 8, 9]. For phylogenetic inference (i.e. to determine the hierarchy of case relationships), sequences were integrated with associated metadata and aligned on a local implementation of NextStrain [10] using augur and displayed via a web browser using auspice.

### Data sharing

Sequence data for viral genomes are deposited in GISAID with the strain names shown in Figure 2 and with the following EPI_ISL accession numbers: 2249220, 2249226, 2376250, 2376251, 2376252, 2500993, 2500994, 2500995, 2500996, 2500997, 2500998 and 2500999.

### Ethical and institutional approval

Specimens were analyzed under a protocol approved by the Gundersen Health System Institutional Review Board (#2-20-03-008; PI: Kenny) to perform next-generation sequencing on remnant specimens after completion of diagnostic testing. Testing of identified specimens was explicitly permitted, as was chart review to correlate viral genome data with data abstracted from clinical notes on diagnosis, symptoms, relevant co-morbidities, clinical course and resolution of the SARS-CoV-2 infection in these patients. Scientific publication of deidentified data was also permitted. This manuscript was additionally reviewed by the Legal Department of Gundersen Health System prior to submission.

### Rapid PCR Test for Outbreak Strain

The strain-defining G19086T polymorphism introduced an ApoI restriction site absent in the reference viral genome. A 319 bp region spanning the G19086T polymorphism was amplified using AATTCCCAGTTCTTCACGACA and AAAGCTGGTGTGTGGAATGC. PCR products were incubated with ApoI and visualized following gel electrophoresis in order to determine the presence of the ApoI site at 19086. All candidate positive specimens were immediately screened for G19086T to facilitate the outbreak investigation, and later confirmed by whole viral genome sequencing.

### Statistical Analysis

Patients and staff who were partially vaccinated or who refused SARS-CoV-2 testing were excluded from the statistical analysis of viral attack rate and vaccine effectiveness, as were non-staff patient caregivers. Uniform viral exposure was assumed for all individuals included in the analysis. Fisher’s exact test was used to compare viral attack rates between groups, and a *p*-value threshold of *p* < 0.05 was set to determine statistical significance. All statistical analysis was performed using the SAS software suite, version 9.4 (SAS Foundation, Cary NC).

## RESULTS

### Outbreak Case Distribution

Dialysis patients generally follow a rigid treatment schedule consisting of three treatments weekly. Due to this, they tend to receive treatment with the same patient cohort on the same days of the week. There are two treatment cohorts at the ESRD facility: one receiving treatment on a schedule “A” and the other treated on schedule “B”. Of the 106 patients who received dialysis in the facility at the time of the outbreak, 10 (P1-P10) tested positive for SARS-CoV-2 infection (9.5%) over a 13-day period, with cases detected among patients attending both of the alternate day dialysis schedules. 3 additional cases of SARS-CoV-2 infection were confirmed among dialysis staff (S1-S3; 6.4%). One associated individual (P11), a patient’s caregiver frequently present at dialysis, also tested positive. Because of the overlapping exposure risk for P11, they are included in the genomic analysis but excluded from the vaccine effectiveness analysis and from other analyses specific to ESRD patients (e.g. hospitalization rate). Case characteristics are presented in detail in Table 2. The outbreak timeline is shown in Figure 1. Nine ESRD patients with infection were symptomatic (90%). Two of the three staff were asymptomatic at the time of testing but indicated experiencing symptoms in the weeks leading up to the cluster that they had attributed to other illnesses. Of the 6 patients requiring hospitalization, only one was fully vaccinated. One unvaccinated patient in the outbreak died following hospitalization. Among the fully vaccinated patients who tested positive, the average time post-vaccine series completion was 64 days (range 44 – 119 days).

**Table 2.** Details of patients and staff who tested positive for SARS-CoV-2 during the outbreak.

| Identifier | Symptom Presentation | Vaccination Status* | Time between most recent vaccination and diagnosis date (weeks) | Hospitalization Status | Patient Died |
| --- | --- | --- | --- | --- | --- |
| P1 | Symptomatic | Fully vaccinated | 6 | - | - |
| P2 | Symptomatic | Partially vaccinated | 2 | Yes | - |
| S1 | Symptomatic | Unvaccinated |  | - | - |
| P3 | Symptomatic | Unvaccinated |  | - | - |
| P4 | Symptomatic | Unvaccinated |  | Yes | Yes |
| P5 | Symptomatic | Unvaccinated |  | Yes | - |
| P6 | Symptomatic | Unvaccinated |  | Yes | - |
| P7 | Symptomatic | Fully vaccinated | 4 | Yes | - |
| P8 | Symptomatic | Unvaccinated |  | - | - |
| S2 | Asymptomatic | Unvaccinated |  | - | - |
| S3 | Asymptomatic | Fully vaccinated | 17 | - | - |
| P9 | Symptomatic | Fully vaccinated | 9 | - | - |
| P10 | Asymptomatic | Fully vaccinated | 8 | - | - |
| P11 | Symptomatic | Unvaccinated |  | - | - |

\*At the time of the outbreak, "Fully vaccinated" was considered two doses either Pfizer or Moderna mRNA vaccine or a single dose of the Johnson & Johnson vaccine.

**Fig 1.**
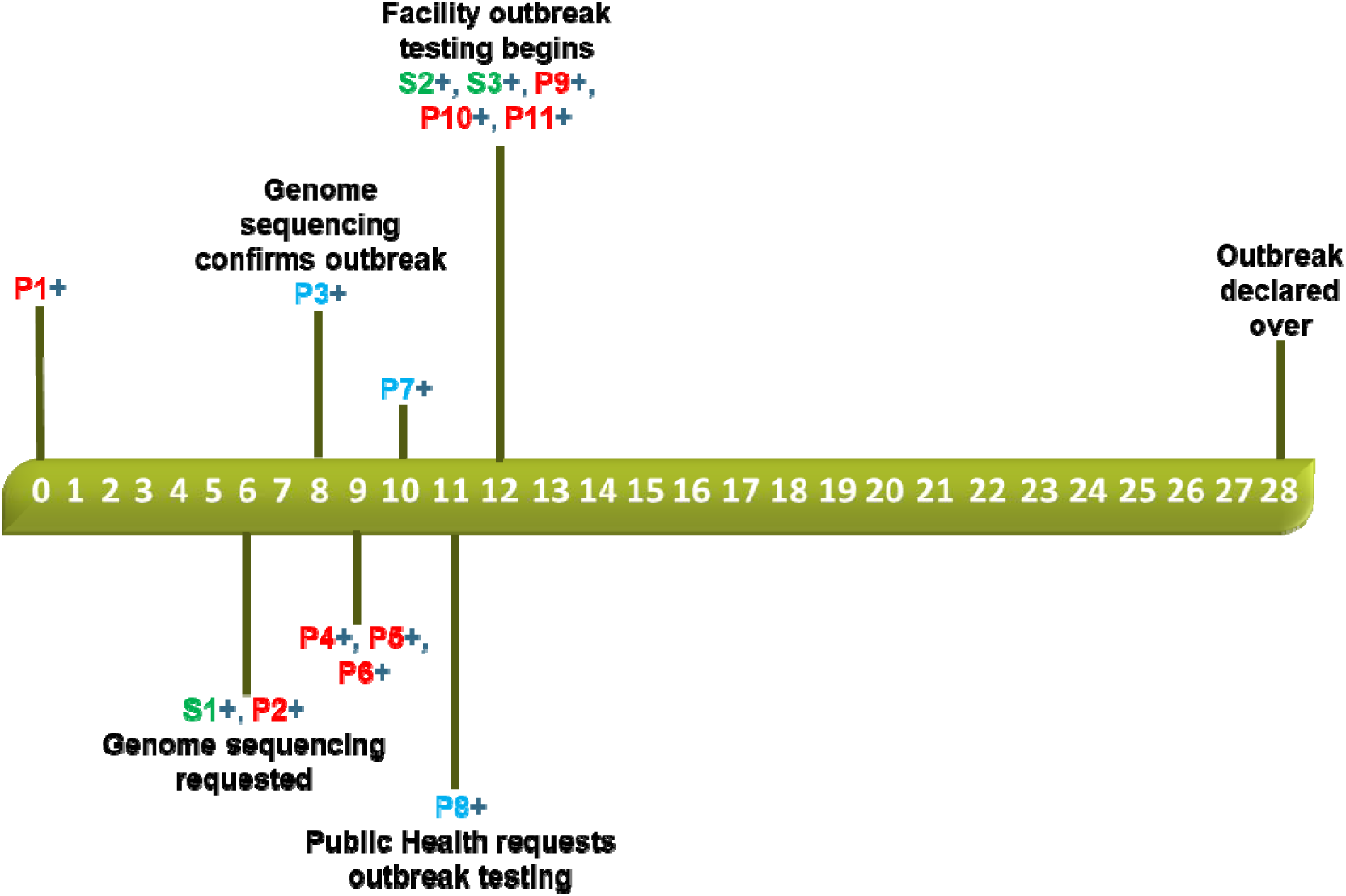
Outbreak Timeline. Cases are identified by an ID (P = Patient, S = Staff), and color-coded by which of two non-overlapping dialysis schedules was utilized by patients (Blue = A, Red = B). Staff cases are indicated in Green.

Viral genome sequencing was requested on Day 6 when the 2^nd^ and 3^rd^ cases were identified and was ultimately performed on 12 of the 14 samples. Two patient samples were tested using methods that did not leave a residual specimen for sequencing (a rapid PCR and a Binax Antigen test). The first two patients (P1 and P2) where infected with the viral strain that had the “S-gene drop-out” by PCR, strongly indicative of the B.1.1.7 substrain, but at the time 64% of regional cases were B.1.1.7. These initial findings were not sufficient to confirm the outbreak. Genetic identity between the first specimens was confirmed by sequencing on day 8, and B.1.1.7/”Alpha” substrain confirmed at that time. This particular B.1.1.7 substrain contained a unique polymorphism (G19086T) which allowed the research team to design a rapid test for the specific outbreak strain. Rapid test validation was completed on Day 13 and was then used to pre-screen all subsequent possible cases prior to sequencing, providing a much more rapid data turnaround to the Infection Control team. Genetic sequencing unambiguously demonstrated that all the cases in the cluster shared an identical substrain of the virus. All genomes were completely identical except P11 which had one additional genetic variant (Figure 2). This made it impossible to infer directionality of infection between patients/staff solely from the genetic data, with the exception of concluding that no individual in the cluster was infected by P11.

**Figure 2.**
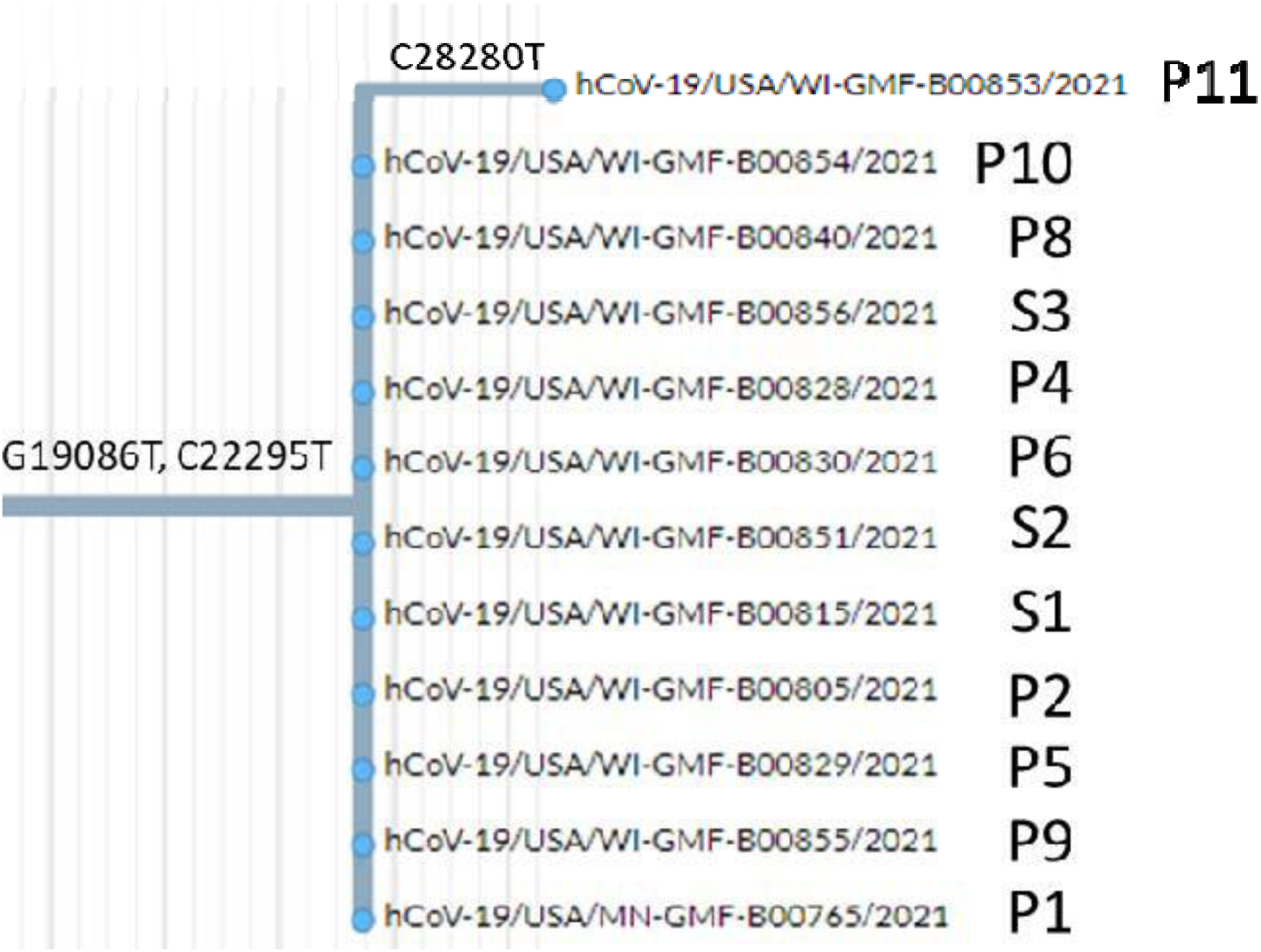
Excerpt of the B.1.1.7 clade from our phylogenetic tree showing the outbreak strain, which was distinguished by two sequence variants from other cases we had sequenced. Specimens aligned vertically are genetically identical.

### Outbreak Epidemiology and Facility Surveillance

The Infection Control department partnered with local and state public health partners to conduct case interviews and infection prevention assessments. The dialysis facility is composed of three treatment pods and operates two dialysis schedules to treat patients on alternate days: schedule A and schedule B. Seven of the ten confirmed patient cases (70%) dialyzed on schedule B. The first patient case (P1, day 0) of the cluster was diagnosed with COVID-19 on a date in the second quarter of 2021 and was determined to have a known community exposure. 6 days after P1 tested positive, two additional individuals (S1, P2) became positive for SARS-CoV-2. Most cases were identified within a 7-day period. The final five cases (P9, P10. P11, S2, S3) were identified during the first week of facility-wide SARS-CoV-2 testing. No additional cases were recorded in weeks two and three of facility testing, indicating the successful control of the outbreak.

A review of the facility’s schedule and staff assignments indicated that two unvaccinated staff members, S1 and S2, cared for 6 and 4 patients in the cluster, respectively. S3 cared for all the patients in the facility though the interactions were for shorter periods of time compared to S1 and S2.

Staff conversations revealed that three affected patients were known to congregate outdoors after dialysis treatment while waiting for transportation, often without masks, and via third-party transportation companies which had limited enforcement of masking standards. Two of these patients transported together to and from treatment. Patient interviews revealed that mask compliance was low during these rides and may have served as a transmission pathway. All three were part of the schedule B cohort, and two of them dialyzed in the same pod at the same time. The clinic vestibule was another noted potential route of transmission when an IPC assessment indicated that this area was not actively monitored for social distancing compliance and patients were lingering there while waiting for rides.

### Infection Prevention Assessment and Interventions

In the spring of 2020, ESRD leadership developed dialysis-specific infection prevention guidelines for SARS-CoV-2. These guidelines included expectations around patient masking, screening, physical distancing, education, and caring for SARS-CoV-2 positive patients.

Education on the importance of hand hygiene and respiratory protection was provided to patients. Vaccination education was added in 2021 following the Food and Drug Administration (FDA) emergency use authorization of COVID-19 vaccines. A summary of our assessment of the effectiveness of these practices at the time of the outbreak is provided in Table 3, along with a description of interventions we implemented while attempting to control this outbreak and reduce the chance of future outbreaks.

**Table 3.** Infection Control Assessments and Interventions.

| **Category** | **Assessment** | **Intervention** |
| --- | --- | --- |
| *A. Masking* | Patients were required to wear a face mask or covering while in the facility. Non-compliant patients were given verbal warnings. If a patient continued to refuse, their dialysis treatment was terminated, and they were sent home. While patients were generally very compliant with masking within the facility, patients were often observed conversing with one another without masks outside of the facility while waiting for transportation, wherein mask adherence was inconsistent. | Patient education was frequently given out to re-emphasize the importance of masking in the prevention of SARS-CoV-2 infection. |
| *B. Symptom Screening-Patients* | Patients were screened for SARS-CoV-2 symptoms and fever when they arrived at the facility. Patients who screened symptomatic during check-in were directed to an “ill-waiting room” where they would be assessed by an RN. All patients underwent a second nursing assessment at chairside prior to the initiation of dialysis. If a patient became symptomatic during treatment, a SARS-CoV-2 RT-PCR test was collected at chairside. Patients were educated that they were to notify the unit if they developed COVID-19-like symptoms.<br>Interviews with staff revealed that patients had the tendency to not disclose their symptoms to screening staff at the entrance of the facility. It wasn’t until they were in the dialysis chair and had a nursing assessment that patients disclosed symptoms such as a cough or unexplained fatigue. Many of the ESRD patients in the facility are medically complex which also disguised SARS-CoV-2 symptoms for some patients. | Screening for symptoms and known exposures is a moderately effective intervention with well-understood limitations [20], but which can still contribute positively as one component of an overall facility strategy. A lack of candor about symptoms and/or exposure [17, 21] is just one of several reasons for screening failures. Recognizing this, no changes were made to the entrance patient screening process but treating staff remained diligent in asking patients about new symptoms and testing accordingly. |
| *C. Symptom Screening-Staff* | Staff were expected to self-screen for symptoms at home and report any new symptoms to the Employee Health department for evaluation and SARS-CoV-2 testing.<br>Two of the three staff that tested positive for SARS-CoV-2 during the outbreak had attributed their symptoms to other causes such as sinus infection and allergies. These symptoms were not reported to Employee Health and these cases were identified during the first week of facility surveillance. | ESRD leadership re-emphasized the organization’s Employee Health policy on SARS-CoV-2 and the importance of reporting new symptoms to the Employee Health Department. |
| *D. Social Distancing and Visitation Policy* | Seating in the waiting room was spaced out to achieve physical distancing. The facility’s visitor policy was also revised by restricting guests with exceptions granted on a case-by-case basis by ESRD leadership. The one conference room in the facility was converted into a second staff breakroom for staff to support physical distancing.<br>No gaps were identified with the facility’s visitation policy or with staff while at work. Major gaps were identified with patient distancing before and after treatment. While the waiting room was constantly monitored, the clinic’s vestibule was not and did contain seating. Patients were also observed sharing benches outside of the clinic while waiting for transportation. | To limit patient congregation in the clinic vestibule, seating was removed from this space and physical distancing signs were posted at the entrance. |
| *E. Caring for SARS-CoV-2 Positive Patients-Isolation Practices* | SARS-CoV-2 positive patients were cared for by dedicated staff in a separate room if available. If a separate room was not available, patients were placed in a treatment chair that promoted physical distancing. Staff wore an isolation gown, eye protection, and respirator (N95 or PAPR, staff choice) throughout the patient's treatment. Dedicated supplies were placed chairside and then disinfected or disposed of after treatment. Following organizational policy, SARS-CoV-2 positive patients were cared for in this manner for 10 days following the positive test. | All SARS-CoV-2 positive and symptomatic patients were cohorted in a designated pod during treatment. These patients were moved to the same afternoon dialysis schedule during treatment and cared for by dedicated staff and supplies. In place of the standard dialysis gown, staff in the COVID cohort group wore a yellow isolation gown to differentiate them from other staff members. The treatment pod also offered the advantage of providing an alternative entry directly into the unit from the parking lot that bypassed the waiting room. Staff called patients once they arrived to admit them into the facility. |
| *F. Personal Protective Equipment (PPE)* | Organizational policy required all staff members to wear a medical grade mask and eye protection when caring for patients in addition to the dialysis-required jacket and gloves. When caring for patients with respiratory symptoms or SARS-CoV-2 positive patients, staff members wore an isolation gown and respirator (N95 or PAPR: staff choice) in addition to standard hemodialysis PPE. During the Infection Prevention Assessment, no gaps were identified with masking, gown, glove, and respirator use. However, compliance with eye protection was variable. Interviews also indicated that staff were not routinely disinfecting their eyewear. | Education was developed on how and when to clean eyewear. ESRD leadership reviewed the importance of regular eyewear disinfection with staff. Staff caring for SARS-CoV-2 patients wore an isolation gown instead of the dialysis jacket to differentiate them from other staff. |
| *G. Ventilation* | The facilities team assessed the unit air exchange rate which is the recommended air exchanges occurring in a space per hour (ACH). This should be a minimum of 6 ACH in patient care areas. The initial ACH rate in the unit was determined to 3.8 ACH. | The facilities team increased the number of air exchanges in the treatment area to 6.3 ACH. |
| *H. Infection Prevention Interventions -Patient* | Patients were provided instructions on hand hygiene, respiratory hygiene, masking, and cough etiquette. | Supplemental vaccine and masking education to re-emphasize the importance of both tools in preventing SARS-CoV-2 infection and reducing morbidity. |
| *I. Vaccine education - Staff* |  | Supplemental vaccine education was developed for staff. With new vaccinations and some staffing changes, the proportion of staff that was fully-vaccinated staff increased from 77% to 84% during the span including and immediately following the outbreak. |

### COVID-19 Vaccination Effectiveness Analysis

At the time of the outbreak, vaccination rates among patients and staff at the ESRD facility were generally high, with approximately 88% of all dialysis patients fully vaccinated, while 77% of dialysis staff were fully vaccinated (Table 1). After excluding partially vaccinated patients (N=3) and patient caregivers (N=1), unvaccinated cases (N=7) had a higher attack rate compared to their fully vaccinated counterparts (33% vs 4% respectively; vaccine effectiveness = 88%; p <0.001) [Table 4]. When patients and staff were analyzed separately, a similar association between vaccination status and SARS-COV-2 testing status was noted across all patients (Table 4, effectiveness = 91%, p < 0.001), while a weak but statistically non-significant association was noted for dialysis staff members (Table 4, effectiveness = 85%, p = 0.13).

**Table 4.** Analysis of vaccine effectiveness in patients and staff.

| Comparison | SARS-CoV-2 status | Vaccinated | Unvaccinated | Vaccine Effectiveness (p-value) |
| --- | --- | --- | --- | --- |
| Combined analysis including all patients and staff [a] |  | N = 129 | N = 21 |  |
|  | Negative | 124 (96%) | 14 (67%) | 88% (<0.001) |
|  | Positive | 5 (4%) | 7 (33%) |  |
| Patients only |  | N=93 | N = 10 |  |
|  | Negative | 89 (96%) | 5 (50%) | 91% (<0.001) |
|  | Positive | 4 (4%) | 5 (50%) |  |
| Staff only |  | N = 36 | N = 11 |  |
|  | Negative | 35 (97%) | 9 (82%) | 85% (0.13) |
|  | Positive | 1 (3%) | 2 (18%) |  |
| Schedule B patient cohort |  | N = 46 | N = 4 |  |
|  | Negative | 43 (93%) | 1 (25%) | 91% (0.004) |
|  | Positive | 3 (7%) | 3 (75%) |  |

[a] Partially vaccinated individuals are excluded from this analysis

Initial contact tracing suggested that the current outbreak started with a schedule B patient, and early transmission events included other schedule B patients who tended to associate with that initial case. Because of this, it is likely that the schedule B cohort had greater SARS-COV-2 exposure compared to the schedule A cohort. A sub-analysis of only the schedule B cohort (Table 4) suggests similar vaccine effectiveness in this subgroup with presumed higher exposure (effectiveness = 91%, p = 0.004).

## DISCUSSION

Our study highlights the ongoing challenges for infection control practice in dialysis centers that persist well into the SARS-CoV-2 vaccine era. Despite substantially higher vaccination percentages among both patients and staff than the averages in comparable dialysis facilities in Wisconsin, and in the midst of ongoing infection control measures at dialysis facilities, the ESRD patients nonetheless remained vulnerable to a significant outbreak event.

Epidemiological investigation and genetic analysis conclusively demonstrated that SARS-CoV-2 transmission was associated with use of this ESRD facility. The facility leadership, research team, and infection control teams partnered to identify potential avenues of exposure, which led to timely reinvigoration of infection prevention interventions and thus brought an abrupt end to the outbreak. Synergism of robust, optimized infection control practices alongside vaccinal coverage among patients and staff alike maximizes the potential of both interventions to decisively terminate outbreaks in progress.

In general, two vaccine doses proved highly effective against morbidity and mortality in this vulnerable population, the sole exception requiring hospitalization being a vaccinated patient receiving immunosuppressive therapy (a setting in which vaccine effectiveness is known to be suboptimal). All other hospitalizations, and the single death in this outbreak, were among unvaccinated individuals. To that end, the described efficacy of the vaccine to avoid the most severe complications of COVID-19 in the general population was also seen in our ESRD population.

Approximately a year and a half into the SARS-CoV-2 pandemic, fatigue to public health recommendations was widely noted among the public [11]. Healthcare workers nationwide were additionally burdened by continual stress [12]. We also found this to be true among both our patients and staff alike. Compounding this fatigue, in the weeks preceding this outbreak there was a succession of guideline amendments eliminating masking outdoors [13], followed closely by dropping masking recommendations for vaccinated individuals in indoor settings [14]. A lack of familiarity with the dynamic state of the evidence regarding vaccine response in ESRD patients [3, 4] may have led to overconfidence in both staff and patients in the prevailing protective measures then still in force. Simultaneously, the profound skepticism among the electively non-vaccinated about the value of masking [15], some genuine confusion about the rapidly changing CDC guidelines [16], and a high prevalence of dishonesty about SARS-CoV-2-related mitigation behaviors [17] rendered the CDC’s apparent hope that non-vaccinated individuals would continue to comply with masking recommendations somewhat unrealistic. In summary, though the described outbreak occurred at time of low community case rates, it nonetheless occurred in wake of rapidly declining mitigation efforts in the community at large, thereby opening an avenue for outbreak propagation in a vulnerable population confronted with the consequences of this generalized laxity in upholding proven measures.

Our previous analysis of a potential outbreak in this same facility concluded that no intra-facility spread had occurred and that the mitigation strategies employed, augmented by timely genetic data to infer transmission patterns, were robustly protective [5]. Yet the viral substrains involved in that outbreak (B.1.2, B.1.1.464 and B.1.139) were notably less transmissible than the B.1.1.7/Alpha variant which caused the currently described outbreak and which is, in turn, less transmissible than the subsequently emerging Delta and Omicron variants. Thus, even if protective measures were maintained at a consistent level, it is possible that failure to adapt practices to more virulent emerging substrains may represent a missed opportunity for infection control. In turn, propensity for outbreaks in vulnerable populations to ensue despite high (or above average) vaccination rates among cohorted patients or staff underscores the importance of achieving near-complete vaccine coverage by all persons implicated in the chain of transmission.

This 14-person cluster logically resulted from 13 person-to-person transmission events in close succession per the genetic data we obtained. An assessment of the magnitude of potential ESRD infection control failures would require knowing how many, if any, transmissions occurred within the ESRD facility itself. The infection of three healthcare workers by the outbreak strain implicates at least some intra-facility spread and provides an explanation for the detection of identical viral genomes among patients on alternate dialysis schedules since the staff may have served as a bridge for transmission between the schedule cohorts. Moreover, intra-facility mitigation efforts would have been insufficient in face of inadequacies in infection control practices taking place during transportation, commingling of patients outside the facility, vaccine uptake among persons in the ESRD patients’ close orbit, and practical limitations imposed on facilities unable to meet all elements of existing guidelines (such as cohorting, segregating, and distancing dialysis patients in markedly resource-limited settings). With no additional cases identified after the first round of testing, our facility demonstrated the effectiveness of a pod with a separate entrance to control the spread of this strain within the facility (Table 3E). All staff entering the area used full respiratory protection with a PAPR or N95 mask. The team also reengaged the facilities team to assess facility air exchange. Collectively, these variables should be reviewed on an ongoing basis during facility quality assurance and performance improvement meetings. Individual behaviors outside the facility during shared transport (by either patients or staff) or pre-/post-treatment socializing were likely of higher prospective risk compared to those risks existing within the facility, and presumably contributed to the size of the cluster, albeit largely outside the control of facility management.

This outbreak occurred several months after widespread availability of SARS-CoV-2 vaccines, by which time rates of new vaccinations had substantially slowed. Evidence for breakthrough infections among the vaccinated were still mostly anecdotal, and rigorous high-quality studies permitting more robust risk estimation [18] had not yet been reported. The full vaccination rates of patients and staff (88% and 77%, respectively), while above statewide averages (70% and 50%, respectively [6]), were insufficient to prevent the outbreak altogether but almost certainly limited both its extent and individual case mortality. Vaccine mandates for healthcare workers had been initiated at that time in only a tiny minority of US hospitals. In mid-August, our institution imposed a vaccination requirement for all staff and, later still, the federal government initiated a nationwide mandate for healthcare organizations in receipt of Medicare and Medicaid funding [19]. Our study underlines the necessity of such mandates to establish and maintain a safe healthcare environment for provision of care to ESRD patients who, despite receiving appropriate vaccinations, may nonetheless remain at higher risk of breakthrough infections in the right epidemiological setting.

## Data Availability

https://www.gisaid.org/

## ACKNOWLEDGEMENTS

We thank Ana Bardossy, MD, and Shannon Novosad, MD for helpful comments on the manuscript, and Jacqueline Cutts, MPH, RN, BSN for collaboration and support. This work was supported by an Emergent Ventures Fast Grant [grant number 2243] to PK and by the Gundersen Medical Foundation. PK holds the Dr. Jon & Betty Kabara Endowed Chair in Precision Oncology.

